# Production of anti-spike antibodies in response to COVID vaccine in lymphoma patients

**DOI:** 10.1101/2022.03.24.22272883

**Authors:** Alexandra Della Pia, Gee Youn (Geeny) Kim, Andrew Ip, Jaeil Ahn, Yanzhi Liu, Simone Kats, Michael Koropsak, Brittany Lukasik, Anamta Contractor, Krushna Amin, Lakshmi Ayyagari, Charles Zhao, Amolika Gupta, Mark Batistick, Lori A. Leslie, Andre Goy, Tatyana Feldman

## Abstract

Patients with hematologic malignancies have poor outcomes from COVID infection and are less likely to mount an antibody response after COVID infection. There is limited data on the efficacy of the COVID vaccines in lymphoma patients, and to suggest the optimal timing of vaccination to elicit immunity in patients receiving immunochemotherapy. This is a retrospective study of adult lymphoma patients who received the COVID vaccine between 12/1/2020 and 11/30/2021. The primary endpoint was a positive anti-COVID spike protein antibody titer following the primary COVID vaccination series. The primary series was defined as 2 doses of the COVID mRNA vaccines or 1 dose of the COVID adenovirus vaccine. Subgroups were compared using Fisher’s exact test, and unadjusted and adjusted logistic regression models were used for univariate (UVA) and multivariate (MVA) analyses. A total of 243 patients were included in this study; 72 patients (30%) with indolent lymphomas; 56 patients (23%) with Burkitt’s, diffuse large B-cell lymphoma (DLBCL), and primary mediastinal B-cell lymphoma (PMBL) combined; 55 patients (22%) with chronic lymphocytic leukemia or small lymphocytic lymphoma (CLL/SLL); and 44 patients (18%) with Hodgkin and T-cell lymphomas (HL/TCL) combined. One-hundred fifty-eight patients (65%) developed anti-COVID spike protein antibodies after completing the primary COVID vaccination series. Thirty-eight of 46 (83%) patients who received an additional primary shot and had resultant levels produced anti-COVID spike protein antibodies. When compared to other lymphoma types, patients with CLL/SLL had a numerically lower seroconversion rate of 51% following the primary series whereas patients with HL/TCL appeared to have a robust antibody response with a seropositivity rate of 77% (p=0.04). Lymphoma patients are capable of mounting a humoral response to the COVID mRNA vaccines. Further studies are required to confirm our findings, including whether T-cell immunity would be of clinical relevance in this patient population.

## Introduction

Patients with hematologic malignancies have poor outcomes from COVID infection with associated mortality of up to 30-40%. Studies have shown that these patients are less likely to mount an antibody response after COVID infection [1]. The Pfizer-BioNTech and Moderna COVID mRNA vaccines have been shown to be 94% effective in preventing severe disease in the general population [2]. However, it is well known that certain clinically vulnerable populations do not develop protective immunity after completing the COVID vaccine series [3].

Production of antibodies against the SARS-CoV-2 spike protein is the major mechanism of protective immunity induced by COVID vaccination. Patients with lymphoma may be unable to seroconvert following vaccination due to impaired humoral and T-cell immunity secondary to the malignancy itself as well as immunosuppressive treatment [4]. This inability to produce antibodies against the SARS-CoV-2 spike protein is most notable in patients with B-cell lymphomas and chronic lymphocytic leukemia (CLL), with seroconversion rates ranging between 64% to 78% [5]. Treatment with B-cell directed therapies, such as anti-CD20 monoclonal antibodies and Bruton’s tyrosine kinase (BTK) inhibitors, further compromises the ability of patients with lymphoma to mount an antibody response following vaccination and persists over time [6]. Vaccine responses in patients with different lymphoma subtypes or receiving B-cell depleting therapies have not been fully elucidated.

With the emergence of the omicron variant and availability of an additional primary shot of the COVID vaccine, it has become increasingly important to describe the efficacy of the COVID vaccines in lymphoma patients and to suggest the optimal timing of vaccination to elicit immunity in patients receiving immunochemotherapy. There have been several reports of patients with lymphoma frequently not achieving serologic response to COVID vaccines, especially after recent treatments with B-cell directed therapies [6,7]. In this study, we evaluated antibody levels against the SARS-CoV-2 spike protein in lymphoma patients following receipt of the primary COVID vaccination series and additional primary shot. We hypothesized that patients with lymphoma are not developing a robust immune response following vaccination.

## Methods

### Study Design and Cohort Selection

This was a retrospective, single center study at the John Theurer Cancer Center (JTCC) within Hackensack Meridian Health (HMH). Electronic health record (EHR)-derived data was utilized to identify patients who received at least 2 doses of the COVID mRNA vaccines or at least 1 dose of the COVID adenovirus vaccine. The primary objective was to evaluate the efficacy of the COVID vaccines following 2 doses of the COVID mRNA vaccines or 1 dose of the COVID adenovirus vaccine.

Patients were included in this analysis if they were: 1) Adult patients ≥18 years; 2) Had a diagnosis of lymphoma or CLL; 3) Received the COVID vaccine series between December 15, 2020 and November 30, 2021; 4) had a resulted anti-COVID spike protein antibody titer following 2 doses of the COVID mRNA vaccines (Pfizer-BioNTech vaccine and/or Moderna COVID mRNA vaccine) or after 1 dose of the COVID adenovirus vaccine (Johnson&Johnson).

Institutional Review Board (IRB) approval was obtained. The requirement for patient informed consent was waived by the IRB as this project represented a non-interventional study utilizing routinely collected data for secondary research purposes.

### Data Sources

Data was collected from HMH’s EHR (Epic) which is utilized throughout the network. Patients treated at JTCC were identified by convenience sampling if they shared their vaccination status at the time of their outpatient visit. Demographics, clinical characteristics, treatments, and outcomes were manually extracted by physicians, pharmacists, nurses, and medical students from JTCC and HMH. Data abstracted by the team was entered into an Excel spreadsheet, and quality control was performed by physicians overseeing data collection.

Demographic data included age, sex, race/ethnicity, co-morbidities, and last anticancer treatment received. Co-morbidities were defined as any of the following present prior to receipt of the COVID vaccine series: chronic obstructive pulmonary disorder (COPD), autoimmune diseases, liver disease, diabetes mellitus, chronic kidney disease (CKD), human immunodeficiency virus (HIV), cardiovascular disease, and smoking history. Lymphoma types were categorized as follows: chronic lymphocytic leukemia (CLL)/small lymphocytic leukemia (SLL); Burkitt’s lymphoma/diffuse large B-cell lymphoma (DLBCL)/primary mediastinal B-cell lymphoma (PMBL); indolent B-cell lymphomas; mantle cell lymphoma (MCL); and Hodgkin’s lymphoma/T-cell lymphomas (HL/TCL). Indolent lymphomas included follicular lymphoma, marginal zone lymphoma, lymphoplasmacytic lymphoma, and Waldenstrom macroglobulinemia. T-cell lymphomas included peripheral T-cell lymphomas not otherwise specified (NOS), anaplastic large cell lymphoma (ALCL), cutaneous T-cell lymphomas, angioimmunoblastic T-cell lymphoma (AITL), and acute T-cell leukemia/lymphoma (ATLL). Most recent anticancer treatment types (such as cytotoxic chemotherapy, anti-CD20 monoclonal antibody therapy, or small molecule inhibitors), date of last treatment prior to vaccination, and duration of treatment were also collected.

### Outcome Measures

The primary outcome was a positive anti-COVID spike protein antibody titer following the primary COVID vaccination series. The primary series was defined as either 2 doses of the COVID mRNA vaccines or 1 dose of the COVID adenovirus vaccine. Among patients who had anti-COVID spike protein antibody results, COVID-19 history was collected and included the following (if available): type of COVID vaccine received and number of doses, history of positive COVID infection, and therapies received for COVID treatment (if applicable).

Secondary outcomes included positive anti-COVID spike protein antibody titers in the following patient subgroups: patients who received an additional primary shot; different lymphoma subtypes; patients who received anti-CD20 monoclonal antibody therapy within the previous 12 months; and patients who received active anticancer treatment. The additional primary shot was defined as either an additional dose (“third dose”) of the COVID mRNA vaccine or an additional dose (“second dose”) of the COVID adenovirus vaccine. Active anticancer treatment was defined as the receipt of any therapy intended to treat cancer that was administered within the six months prior to initiating the primary COVID vaccination series.

### Statistical Analysis

Demographic and clinical characteristics were summarized using descriptive statistics such as percentiles, median, and interquartile range (IQR) by the primary series and additional shot groups, respectively. Seropositivity rates between demographic and clinical categories were compared using Fisher’s exact test or Pearson’s chi-squared tests. Unadjusted and multivariable adjusted logistic regression models (UVA and MVA, respectively) using variables with p<0.05 in the unadjusted model were then fitted and their odds ratios (OR) and confidence intervals (CI) were computed. A two-sided p-value <0.05 is considered for the statistical significance. All analyses were performed using R (*ver* 4.1)

## Results

### Patient demographics and comorbidities

Two-hundred forty-three patients were identified with baseline characteristics as shown in Table 1. Overall, the study population was elderly at a median age of 67 (IQR 59-75) years old, 52% of patients were male, and 78% of patients were white. The most frequent comorbidities were cardiovascular disease (44%) and former smoking history (30%). Lymphoma subtypes in our cohort were: indolent lymphomas (30%), Burkitt’s, DLBCL, PMBL combined (23%), CLL/SLL (22%), and Hodgkin’s and T-cell lymphomas (HL/TCL) combined (18%). Of patients who completed their primary vaccination series, one-hundred twelve (46%) patients received an additional primary shot as of November 30, 2021.

**Table 1.** Baseline characteristics.

| Patient characteristics | Primary series<br>N=243<br>n(%) | Additional primary shot<br>N=112<br>n(%) |
| --- | --- | --- |
| Age, years, median (IQR) | 67 (59-75) | 67 (59-75) |
| Male | 127 (52) | 59 (53) |
| **Race** |  |  |
| Asian | 4 (2) | 0 |
| Black or African American | 11 (4) | 3 (3) |
| Hispanic or Latino | 23 (10) | 6 (5) |
| White | 189 (78) | 98 (88) |
| Other | 11 (4) | 3 (3) |
| Missing | 5 (2) | 2 (2) |
| **Disease type** |  |  |
| Burkitt's, DLBCL, PMBL | 56 (23) | 24 (21) |
| CLL/SLL | 55 (22) | 24 (21) |
| Hodgkin's lymphomas (HL) | 20 (8) | 9 (8) |
| Indolent lymphomas<sup>†</sup> | 72 (30) | 36 (32) |
| Mantle cell lymphoma (MCL) | 12 (5) | 8 (7) |
| T-cell lymphomas (TCL)<sup>‡</sup> | 24 (10) | 8 (7) |
| Other<sup>§</sup> | 4 (2) | 3 (3) |
| **Comorbidities** |  |  |
| *Cardiovascular disease*<sup>¶</sup> | 106 (44) | 49 (44) |
| *COPD* | 6 (3) | 3 (3) |
| *Autoimmune disease*<sup>††</sup> | 27 (11) | 13 (12) |
| *Mild liver disease*<sup>‡‡</sup> | 6 (3) | 2 (2) |
| *Diabetes* | 38 (16) | 19 (17) |
| *Chronic kidney disease* | 10 (4) | 7 (6) |
| *HIV/AIDS* | 6 (3) | 2 (2) |
| *Smoking history* |  |  |
| Current smoker | 8 (3) | 4 (4) |
| Former smoker | 74 (30) | 33 (30) |
| **Vaccine received** |  |  |
| *Pfizer-BioNTech* | 115 (47) | 66 (59) |
| *Moderna* | 79 (33) | 41 (37) |
| *Johnson&Johnson* | 3 (1) | 0 |
| *Unknown* | 46 (19) | 5 (4) |
| <sup>†</sup>Includes follicular lymphomas, marginal zone lymphoma, lymphoplasmacytic lymphoma, and Waldenstrom macroglobulinemia<br><sup>‡</sup>Includes peripheral T-cell lymphomas not otherwise specified (NOS), anaplastic large cell lymphoma (ALCL), cutaneous T-cell lymphomas, angioimmunoblastic T-cell lymphoma (AITL), and acute T-cell leukemia/lymphoma (ATLL)<br><sup>§</sup>Include monoclonal gammopathy of undetermined significance (MGUS), T-cell prolymphocytic leukemia (T-PLL), Essential thrombocytosis, and primary CNS lymphoma<br><sup>¶</sup>Cardiovascular disease defined as coronary artery disease, hypertension, or cerebrovascular accident<br><sup>††</sup>Includes autoimmune diseases, rheumatological disorders, and connective tissue disease<br><sup>‡‡</sup>Mild liver disease defined as either chronic hepatitis or cirrhosis without portal hypertension<br>DLBCL=diffuse large B-cell lymphoma; PMBL=primary mediastinal B-cell lymphoma; COPD=chronic obstructive pulmonary disease; HIV/AIDS=human immunodeficiency virus/acquired immunodeficiency syndrome |  |  |

Testing for anti-COVID spike protein antibodies occurred at a median of 61 (IQR 29-98) days after completion of the primary vaccination series and at a median of 37 (IQR 21-56) days after an additional primary shot. Majority of patients received COVID mRNA vaccines, and we were able to confirm the specific type in 197 (81%) patients. Only 3 patients received the COVID adenovirus vaccine (Table 1).

### Primary outcome

One-hundred fifty-eight patients (65%) developed anti-COVID spike protein antibodies after completing their primary COVID vaccination series (Fig 1 and Table 2). There were 112 patients who received an additional primary shot, but only 46 of these patients had anti-COVID spike antibody protein levels resulted at the time of data cut-off. Of these 46 patients, 38 patients (83%) had anti-COVID spike protein antibodies after receiving this dose (Table 2). Among the 38 patients who were seropositive after the additional primary dose, 18 patients were seronegative after their primary COVID vaccination series and seroconverted only after the additional primary shot of COVID vaccine.

**Figure 1.**
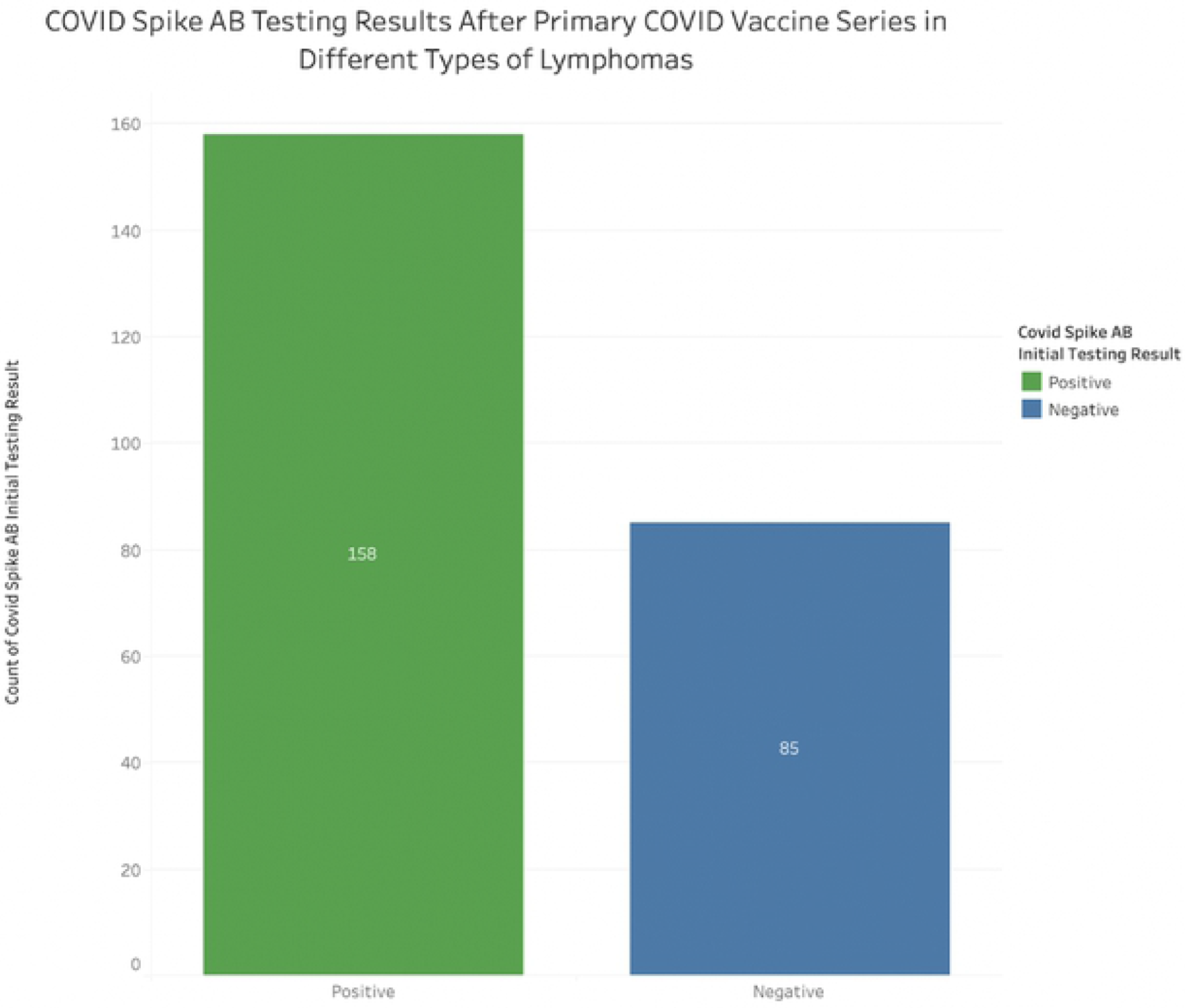
Serologic response against COVID spike protein after the primary series in the overall population.

**Table 2.**
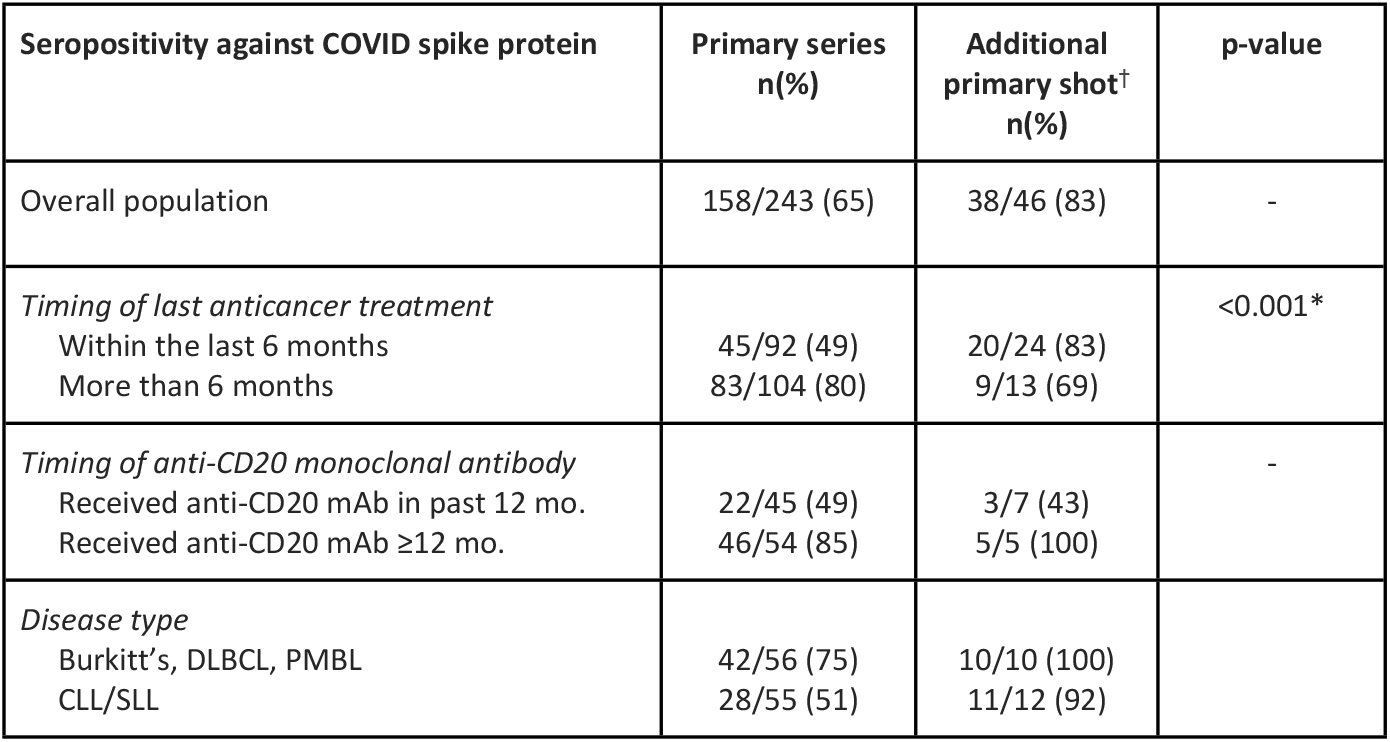
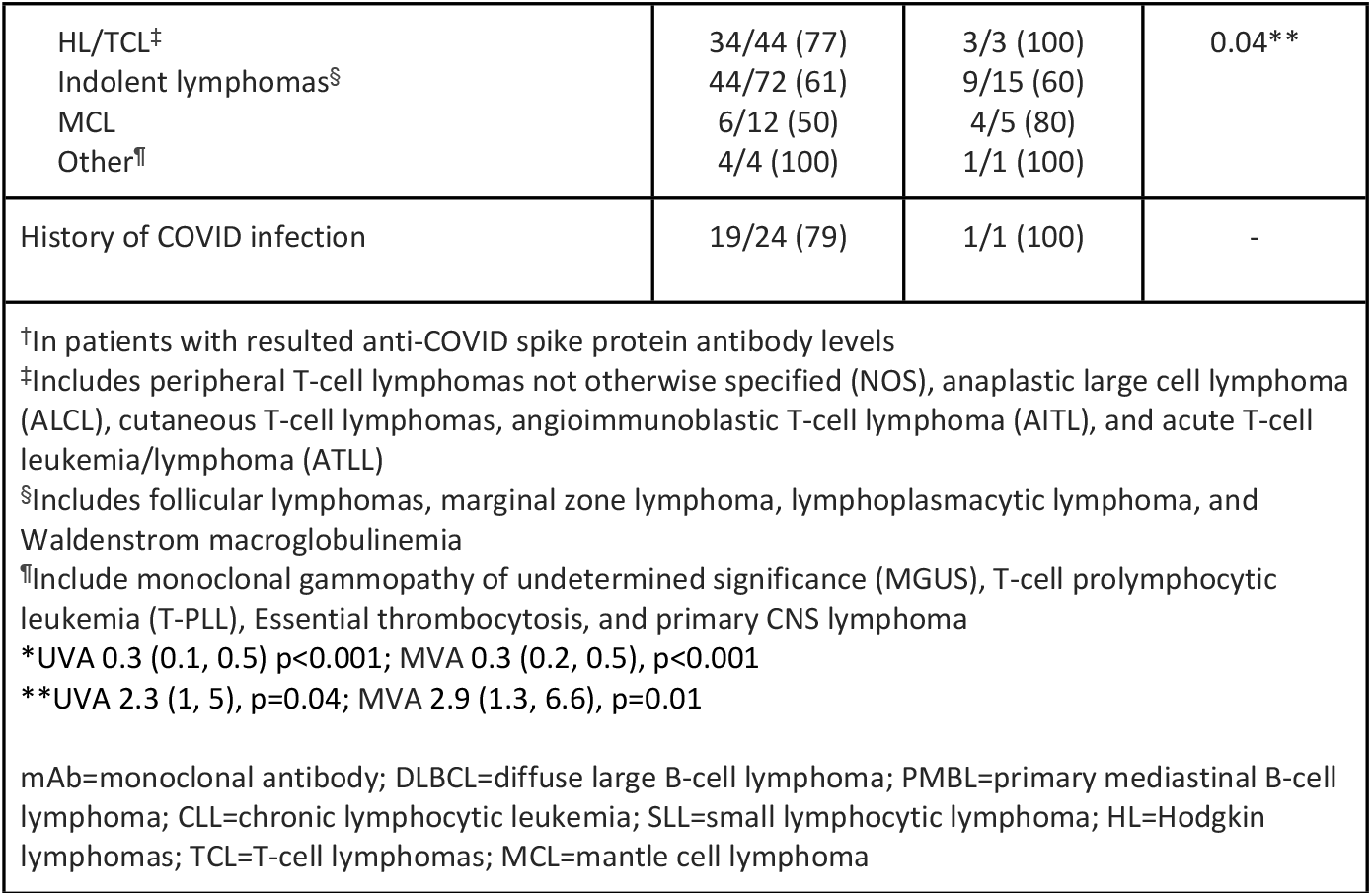
Results.

There were 8 patients that were seronegative following the additional primary shot. Of these 8 patients, there was 1 patient that was seropositive after the primary series but was seronegative after the additional primary shot. There were 7 patients that did not produce antibodies following both the primary vaccination series and additional primary shot. Of these 7 patients, 5 (71%) were diagnosed with an indolent lymphoma, 1 (14%) with CLL/SLL, and 1 (14%) with MCL. At the time of vaccination, 4 of these 7 (57%) patients were receiving active anticancer treatment. Active anticancer treatment in these patients consisted of 50% (2/4) BTK inhibitor monotherapy, 25% (1/4) rituximab with chemotherapy, and 25% (1/4) other therapy.

### Secondary outcomes

There were 45 patients who received an anti-CD20 monoclonal antibody-containing regimen in the last 12 months prior to vaccination, and 22 (49%) of these patients produced antibodies after the primary COVID vaccination series (Table 2). This rate was numerically lower than 85% (46/54) of those who developed antibodies and received an anti-CD20 antibody more than 12 months prior to vaccination. There were 5 patients who received an additional primary shot, of which all (5/5, 100%) were seropositive.

There were differences observed in the ability to produce serology with the COVID vaccine amongst lymphoma subtypes. Of 55 patients with CLL/SLL, 28 (51%) produced antibodies after the primary COVID vaccination series (Table 2 and Fig 2). There were 12 patients who received an additional primary shot, of which 92% were seropositive (Fig 3). There were 20 CLL/SLL patients receiving anticancer treatment at the time of vaccination, of which 7 (7/20, 35%) patients were seropositive after the primary series and 6 (6/7, 86%) after the additional primary shot (Table 3). The most common treatment type in these patients was monotherapy with a Bruton’s tyrosine kinase (BTK) inhibitor, where 36% (4/11) of patients produced antibodies. In comparison, there were 35 CLL/SLL patients not receiving active anticancer treatment and had a numerically higher rate of seroconversion at 60% (21/35) following receipt of the primary COVID vaccine series. CLL/SLL patients were less likely to mount an antibody response to the COVID vaccine when compared to those with other types of lymphoma.

**Figure 2.**
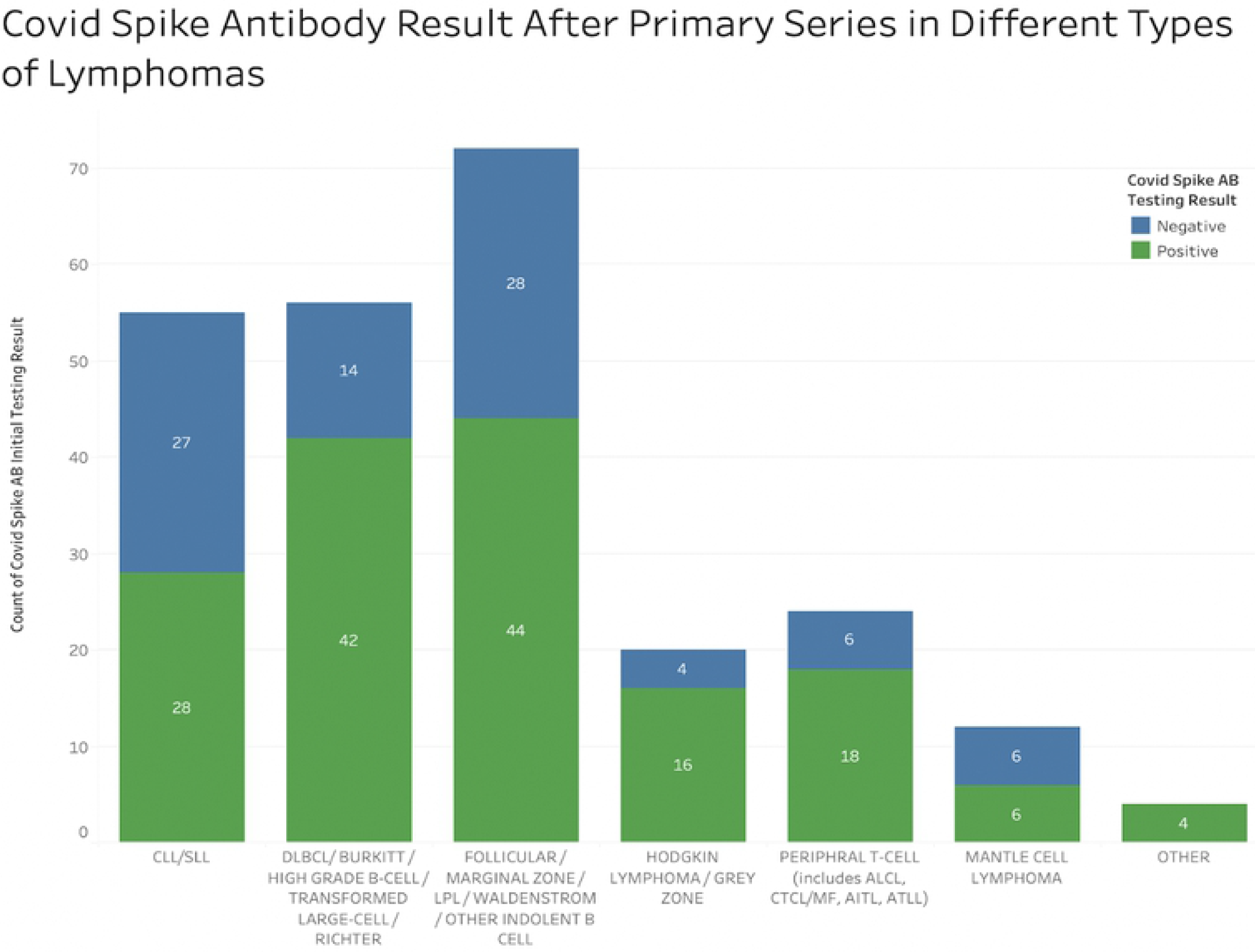
Serologic response against COVID spike protein after primary series by lymphoma diagnosis.

**Figure 3.**
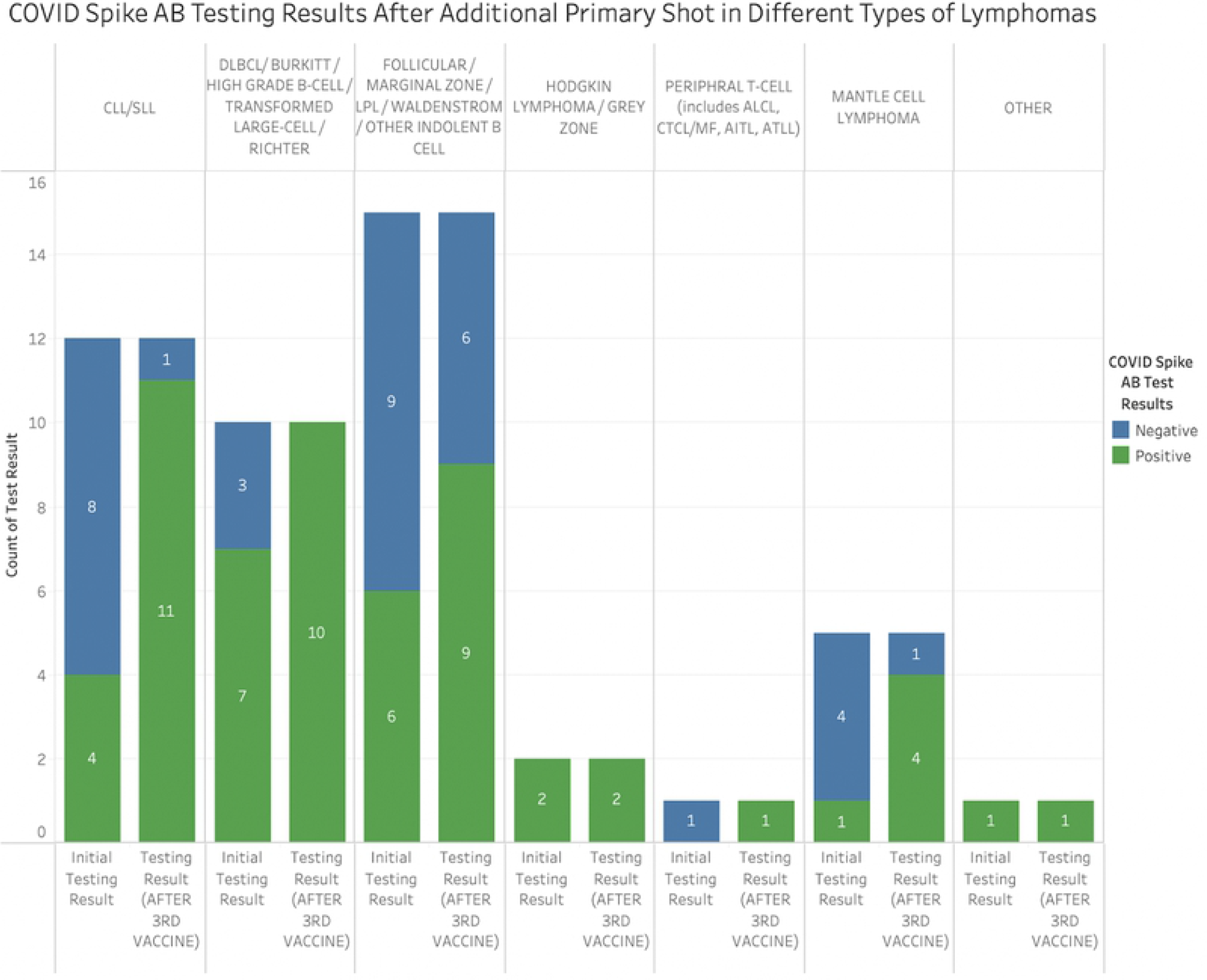
Serologic response against COVID spike protein after additional primary shot by lymphoma diagnosis.

**Table 3.**
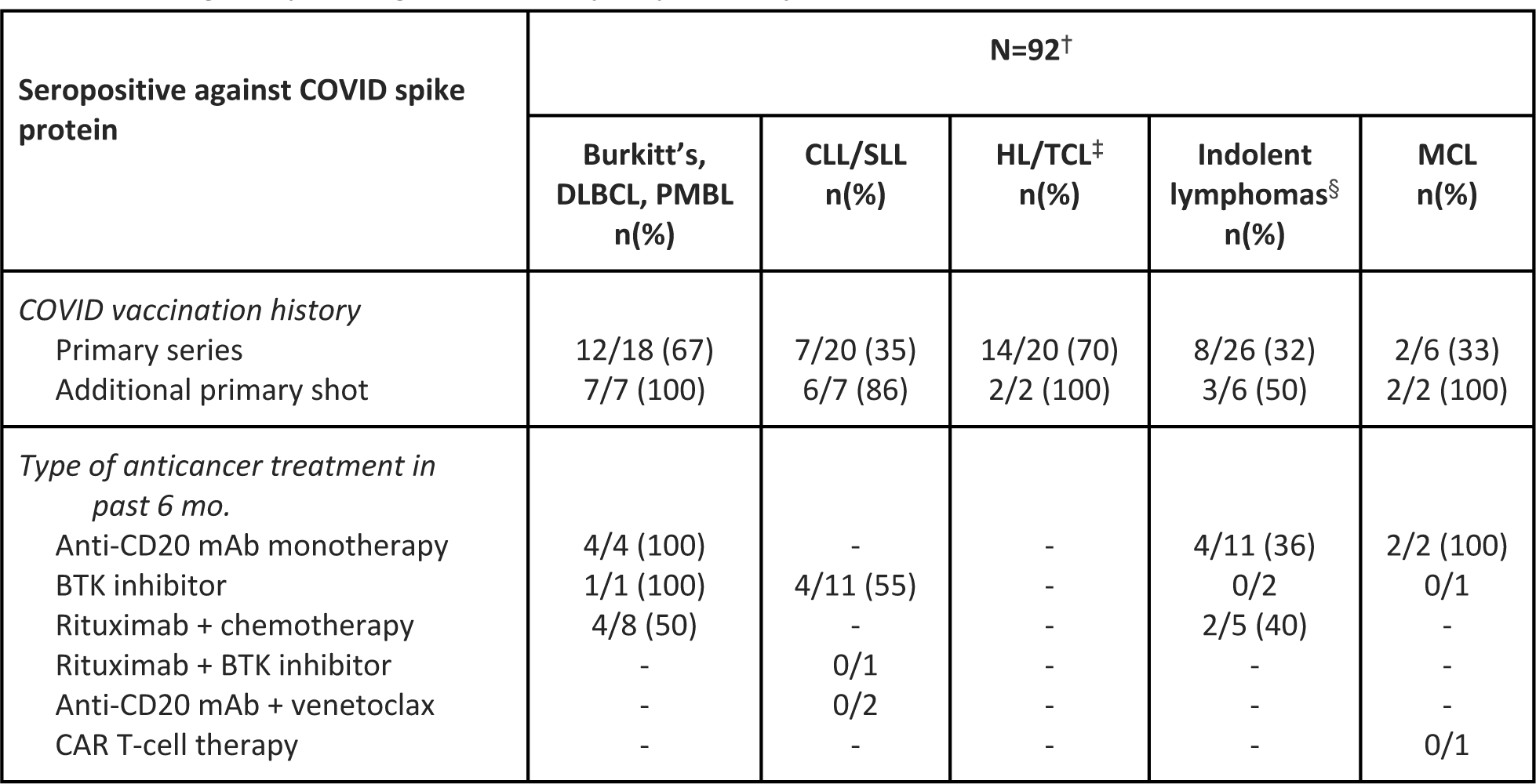
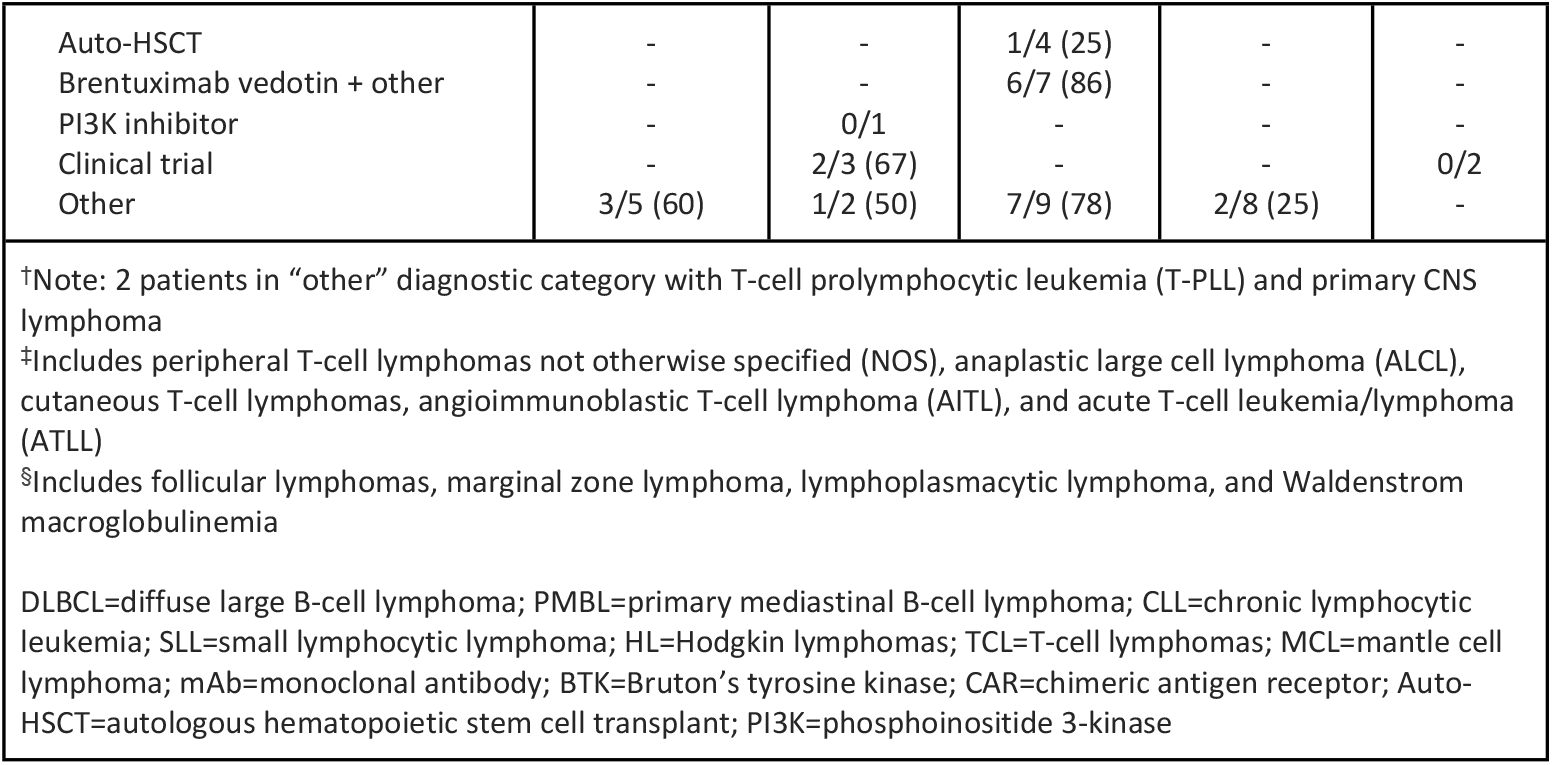
Serologic response against COVID spike protein by active anticancer treatment.

| Seropositive against COVID spike protein | N=92<sup>†</sup> |  |  |  |  |
| --- | --- | --- | --- | --- | --- |
|  | Burkitt's, DLBCL, PMBL n(%) | CLL/SLL n(%) | HL/TCL<sup>‡</sup> n(%) | Indolent lymphomas<sup>§</sup> n(%) | MCL n(%) |
| *COVID vaccination history* |  |  |  |  |  |
| Primary series | 12/18 (67) | 7/20 (35) | 14/20 (70) | 8/26 (32) | 2/6 (33) |
| Additional primary shot | 7/7 (100) | 6/7 (86) | 2/2 (100) | 3/6 (50) | 2/2 (100) |
| *Type of anticancer treatment in past 6 mo.* |  |  |  |  |  |
| Anti-CD20 mAb monotherapy | 4/4 (100) | - | - | 4/11 (36) | 2/2 (100) |
| BTK inhibitor | 1/1 (100) | 4/11 (55) | - | 0/2 | 0/1 |
| Rituximab + chemotherapy | 4/8 (50) | - | - | 2/5 (40) | - |
| Rituximab + BTK inhibitor | - | 0/1 | - | - | - |
| Anti-CD20 mAb + venetoclax | - | 0/2 | - | - | - |
| CAR T-cell therapy | - | - | - | - | 0/1 |
| Auto-HSCT | - | - | 1/4 (25) | - | - |
| Brentuximab vedotin + other | - | - | 6/7 (86) | - | - |
| PI3K inhibitor | - | 0/1 | - | - | - |
| Clinical trial | - | 2/3 (67) | - | - | 0/2 |
| Other | 3/5 (60) | 1/2 (50) | 7/9 (78) | 2/8 (25) | - |

†Note: 2 patients in “other” diagnostic category with T-cell prolymphocytic leukemia (T-PLL) and primary CNS lymphoma ‡Includes peripheral T-cell lymphomas not otherwise specified (NOS), anaplastic large cell lymphoma (ALCL), cutaneous T-cell lymphomas, angioimmunoblastic T-cell lymphoma (AITL), and acute T-cell leukemia/lymphoma (ATLL) §Includes follicular lymphomas, marginal zone lymphoma, lymphoplasmacytic lymphoma, and Waldenstrom macroglobulinemia
DLBCL=diffuse large B-cell lymphoma; PMBL=primary mediastinal B-cell lymphoma; CLL=chronic lymphocytic leukemia; SLL=small lymphocytic lymphoma; HL=Hodgkin lymphomas; TCL=T-cell lymphomas; MCL=mantle cell lymphoma; mAb=monoclonal antibody; BTK=Bruton’s tyrosine kinase; CAR=chimeric antigen receptor; Auto-HSCT=autologous hematopoietic stem cell transplant; PI3K=phosphoinositide 3-kinase

For patients with HL/TCL, 34 of 44 (77%) patients produced antibodies after the primary series and all patients (3/3, 100%) after the additional primary shot (Table 2, Fig 2 and Fig 3). Of 20 HL/TCL patients who received anticancer treatment in the last 6 months, 70% (14/20) produced antibodies at a median titer of 120 AU/mL (reference >=15 AU/mL), with 7 patients having a robust response of antibody titers >400 AU/mL. On statistical analysis, HL/TCL patients were more likely to elicit an antibody response to the COVID vaccine when compared to those with other types of lymphoma, and this response was significant on UVA (OR 2.285, 95% CI (1.038, 5.028), p=0.04) and MVA (OR 2.889, 95% CI (1.260, 6.628), p=0.012) when active anticancer therapy, lymphoma diagnosis, and history of COVID infection were considered in the multivariate analysis.

There were 24 patients who had a documented COVID infection. Of these 24 patients, there were 13 patients who had a positive COVID PCR test result prior to receiving the primary COVID vaccine series; 3 patients had positive COVID PCR test results following completion of the primary series and serology testing; 1 patient had completed the primary vaccine series and had COVID infection prior to serology testing; and 6 patients that were indeterminable due an unknown date of COVID vaccination or COVID PCR test result. In these patients with documented COVID infection, 19 (19/24, 79%) patients were seropositive after the primary vaccination series (Table 2). There were 2 patients who received convalescent plasma, 4 patients who received monoclonal antibody therapy, and 1 patient who received both convalescent plasma and monoclonal antibody treatment for COVID treatment.

## Discussion

In this retrospective, single center study, lymphoma patients (65%) were capable of mounting a humoral response to the primary COVID vaccination series. The majority of patients completed their primary series and additional primary shot with the COVID mRNA vaccines. The ability to produce antibodies to the COVID vaccine appeared to vary according to their lymphoma diagnosis and type of active anticancer treatment received in the past 6 months.

Failure to generate a competent humoral immune response against COVID is associated with an elevated risk of infection and poorer prognosis. Among all patients with hematological malignancies, those with B-cell malignancies generally have suboptimal antibody responses following vaccination against COVID, ranging from 44%-79%, depending on the subtype. This is compared with seropositivity of 91%, 88%, and 95% in vaccinated patients with AML, ALL, and MM, respectively [5]. Among the leukemias, CLL patients have the lowest rate of seroconversion following vaccination, with 64% of patients failing to produce sufficient quantities of anti-spike protein antibodies [5]. The low rate of seroconversion in patients with CLL was also observed in this study. Approximately half (51%) of patients with CLL/SLL produced antibodies following their COVID vaccine, which was considerably lower when compared to most subtypes of lymphoma. In contrast, patients with HL/TCL have been shown to produce antibodies at higher rates 85%-98%, and this may be attributed to differences in disease biology [5]. The results of this study showed patients with HL/TCL had significantly higher rates of seroconversion follow the primary series at 77% compared to other lymphoma subtypes (UVA (OR 2.285, 95% CI (1.038, 5.028), p=0.04) and MVA (OR 2.889, 95% CI (1.260, 6.628), p=0.012).

Seroconversion rates following vaccination are further reduced among patients receiving BTK inhibitors, anti-CD20 monoclonal antibodies, or the BCL2 inhibitor venetoclax [5]. Bruton’s tyrosine kinase is a cytoplasmic tyrosine kinase responsible for B-cell maturation that is constitutively activated in CLL. The CD20 antigen is a pan-B cell marker elevated in certain B-cell lymphomas. BCL2 is an anti-apoptotic protein expressed in high quantities in cancer cells that inhibits the intrinsic pathway of apoptosis. Targeted therapy against these three markers have proven to be effective therapeutic strategies against various B-cell malignancies by suppressing their functionality, including the capacity to generate neutralizing antibodies. In our study, patients with CLL/SLL who received BTK inhibitors with or without rituximab in the past 6 months had suboptimal antibody responses to the COVID vaccine.

For patients receiving B-cell directed therapies, peripheral blood B-cell counts may begin to recover after 3 months following treatment and may be completely reversed by 9 to 12 months [6]. A meta-analysis of thirty-eight studies in patients treated with anti-CD20 monoclonal antibody therapy demonstrated that patients actively on these therapies may have suboptimal antibody responses to all types of vaccines. This phenomenon was determined to be at least partially ameliorated by 3-6 months following treatment, with repopulation of the B-cell pool in 12 months after treatment [8,9]. Similarly, patients in this study who received rituximab in the previous 12 months had numerically lower seropositivity rates to the COVID vaccine compared to patients who had received rituximab more than 12 months prior to vaccination. Of note, the majority of patients with Burkitt’s, DLBCL or PMBL in this study were not actively receiving anticancer therapy in the months prior to COVID vaccination and 75% were able to mount a humoral response to vaccination. This suggests the ability of these patients to recover B-cell counts following B-cell directed therapies and the need for optimal timing of vaccination following these therapies.

Furthermore, this adds to the complexity of predicting response to vaccines in patients with lymphoma. Not only is it affected by the biology of the disease itself, but also the type of therapy they are receiving as well as its timing.

There are some limitations to this study. Firstly, this was a single-center, retrospective chart review. Some patients received their COVID vaccine at an outside facility, therefore, the exact date and the type of vaccine received was not consistently available. Secondly, the timing of the additional primary shot of COVID vaccine was not uniform among all patients, and not all patients received this additional dose. As the COVID-19 pandemic progressed and the Center for Disease Control updated recommendations based on available evidence, some patients received the additional primary shot of the COVID vaccine closer to the recommended 28 days after completion of the primary series than others, and some have yet to receive their additional primary shot. Lastly, although there were 243 patients evaluated in the overall analysis, some subgroups of particular interest had smaller numbers of patients.

In conclusion, lymphoma patients are capable of mounting a humoral response to the COVID mRNA vaccines. Although seroconversion is lower than in the normal population, it is still significant and warrants administration of COVID vaccine without delay, particularly during continuing high rate of infection. CLL/SLL appears predictive of a negative antibody response to the COVID vaccine, while HL/TCL histologies appeared to correlate to a positive antibody response, even with treatment within 6 months of vaccination. Our study suggests anti-CD20 monoclonal antibody therapy in the last 12 months may affect the ability to produce serology towards a COVID vaccine. Further studies are required to confirm our findings, including whether T-cell immunity would be of clinical relevance in this patient population.

## Data Availability

The study involves a review of the electronic medical records of lymphoma patients who received the COVID vaccine. The research data set has removed most, but not all protected health information (for example: actual dates of antibody testing were included as needed for analysis, but would be considered PHI). The dataset is also considered property of Hackensack Meridian Health and not owned by the investigators. Therefore, for both reasons, the dataset cannot be made openly available. However, upon request we are willing to share portions of the data for appropriate review. Requests can be made through the corresponding author or directly to representatives of Hackensack Meridian Health (Dr Andrew Ip).

